# Appraisal and extension of the ERS/ATS Interpretative Strategy for Pulmonary Diffusing Capacity

**DOI:** 10.64898/2026.01.30.26345192

**Authors:** Sylvia Verbanck, Mike Hughes, Femke Demolder, Shauni Wellekens, Stefanie Vincken, Eef Vanderhelst, Shane Hanon

## Abstract

The ERS/ATS22 interpretative flowchart classifies diffusing capacity (DLco) into 5 scenarios with associated pathophysiology, and has not been tested on large patient groups. We aimed to obtain a more layered DLco interpretation, by interrogating DLco components Kco and V_A_, and by estimating lung inflation during the DLco test to identify the presence of restriction, which crucially impacts Kco interpretation. By assessing a “low V_A_” against lung inflation, a novel 9-scenario DLco classification with associated pathophysiology can be obtained. Lung patients from a tertiary center were classified according to the ERS/ATS22 chart and the novel 9-scenario one. Besides a control group of healthy subjects (n=303), disease groups under study were the following : asthma (n=1615), COPD (n=1338), CF (n=108), extrapulmonary restriction (n=122), ILD (n=98), post-COVID (n=193). Except for COPD, the prevalence of “normal DLco” (ERS/ATS22) was generally greater than that of “normal V_A_ and normal Kco” (9-scenario); this discrepancy was most marked in CF (81% *vs* 56%) and in extrapulmonary restriction (57% *vs* 37%). With the novel 9-scenario chart, patients from very different diagnostic groups with a “low DLco” due to emphysema, bronchial disease, interstitial damage or incomplete expansion got classified across distinct scenarios, whereas ERS/ATS22 just grouped them together. In conclusion, when “low V_A_” is evaluated against lung inflation, a differentiation of DLco interpretation can be obtained in various patient groups involving obstruction and/or restriction. This approach can be readily implemented in clinical practice.

## INTRODUCTION

While being pivotal to lung function assessment, the diffusing capacity (DLco) test for carbon monoxide has been challenging to interpret (1,2,3,4), partly due to the respective roles of its two primary constituents, gas transfer coefficient (Kco) and lung volume at which Kco is being measured (alveolar volume, V_A_) (4). In the ATS/ERS2005 guideline (2) predicted DLco behavior was listed for a number of physiological scenarios which might be encountered in patients presenting at the lung clinic. The impact on DLco of abnormalities of pulmonary origin (airway, tissue, vasculature) or of extrapulmonary origin was predicted, based on their respective effects on alveolar membrane and capillary components of diffusion across the air-blood barrier, and on the V_A_ estimate. Neder *et al* (3) further scrutinized DLco behavior with more emphasis on its V_A_ and K_CO_ components, illustrating this with individual clinical cases of COPD or ILD patients. In the most recent ERS/ATS2022 guideline (1), some of these pathophysiological conditions were pooled into 5 DL_CO_ scenarios, where for instance emphysematous and interstitial pathology invariably end up in the same scenarios.

When some degree of restriction is involved, Kco - and thus DLco - become difficult to interpret, as evidenced in the recent past by the challenges of DLco interpretation in COVID patients (5). This is because Kco inherently increases at lower lung inflation (4,6), and therefore it is crucial to establish whether a low V_A_ is, at least partly, due to restriction. This can be done by asking the question: is “low V_A_” due to a reduction in the maximal inflation the patient can achieve during the DLco test (TLC_DLCO_) ? If V_A_ is low and TLC_DLCO_ is low, Kco interpretation specific to a restricted lung applies (4), irrespective of degree of uneven mixing. If V_A_ is low but TLC_DLCO_ is not, this automatically implies that uneven mixing is entirely responsible for the low V_A_, and Kco abnormality can be directly gauged against reference Kco values obtain for healthy subjects at TLC. It is sometimes suggested that if plethysmography were available, V_A_/TLC_pl_ could be used to determine whether uneven mixing is responsible for lowering DLco component V_A_. In this case the question becomes : is a “low V_A_” due to a “low V_A_/TLC” ? A yes or no then merely signals presence or absence of uneven mixing, but does not inform whether any degree of restriction is involved as well. As a result, emphysematous and interstitial pathologies cannot be distinguished at all based on DLco and V_A_/TLC. By combining V_A_, TLC_DLCO_ and Kco, the restrictive component can be identified, obtaining differentiation between 9 distinct scenarios with associated pathophysiology. In this case DLco is merely the result of abnormalities detected in its primary measures V_A_ and Kco, which include cases where underlying V_A_ and Kco abnormalities combine to produce a DL_CO_ within normal limits.

Of the various scenarios described in both 2005 and 2022 documents (1,2) high DLco (and high Kco) was presumed to occur in a significant portion of asthma patients (7). Given the prevalence of asthma, this stimulated us to evaluate the ERS/ATS22 scenarios encountered in our lung function laboratory, not only for asthma patients, but also in COPD, and in other diseases such as cystic fibrosis, muscle weakness, various types of ILD, or post-COVID. The first aim of this study was to classify 3000+ patients with a documented clinical diagnosis, to determine which of the 5 ERS/ATS22 scenarios most frequently occurs within each clinical category. The second aim was to explore whether the patients with lung diseases involving a restrictive component (e.g., ILD) fall into different categories than for instance COPD when also testing V_A_ against lung inflation during the DLco test.

## METHODS

This retrospective study was approved by the UZBrussel ethics committee (B.U.N.1432025000177) and pertained to patients with pneumologist diagnoses including asthma (GINA), COPD (GOLD with grades 1-4), adult cystic fibrosis (Adult CF center), disease of extrapulmonary origin (Neuromuscular reference center), and interstitial lung disease (ILD) (multidisciplinary team). ILD was subdivided into (sarcoidosis II-V, nonspecific interstitial pneumonia (NSIP) and usual interstitial pneumonia (UIP). A separate group of patients previously studied 10 weeks after severe SARS-CoV-2 infection (8) were also retrieved (post-COVID group). Patients from any disease group with BMI>35 kg/m^2^ were excluded. Healthy subjects were retrieved from previous lung function reference value studies (9,10). From our out-patient database, we retrieved only those patients falling within the 20-80 years age range and with full lung function testing. Measurements included DLco testing and plethysmography with quality control described in the Online Supplement; DLco measurements were haemoglobin-corrected.

### Classification of various diseases into 5 or 9 DLco scenarios

The 5-scenario ERS/ATS22 classification chart in Figure 1 is based on Figure 11 in the ERS/ATS22 guideline (1), “*a reasonable interpretation algorithm using DLCO along with KCO and VA*”. Following the indications in that Figure 11, “DLco <LLN” and “DLco >ULN” were readily incorporated by using DLco z-score thresholds at −1.645 and +1.645. The V_A_ indications in the ERS/ATS22 flow chart (“low” or “normal”) were interpreted based on the V_A_ z-score threshold at −1.645, and the Kco indications (“low/normal” or “high”) were interpreted based on the Kco z-score threshold at +1.645. This leads to 5 relevant scenarios (***I***,***II***,***II***,***IV***,***V***) with associated pathologies, as shown in Figure 1.

**Figure 1:**
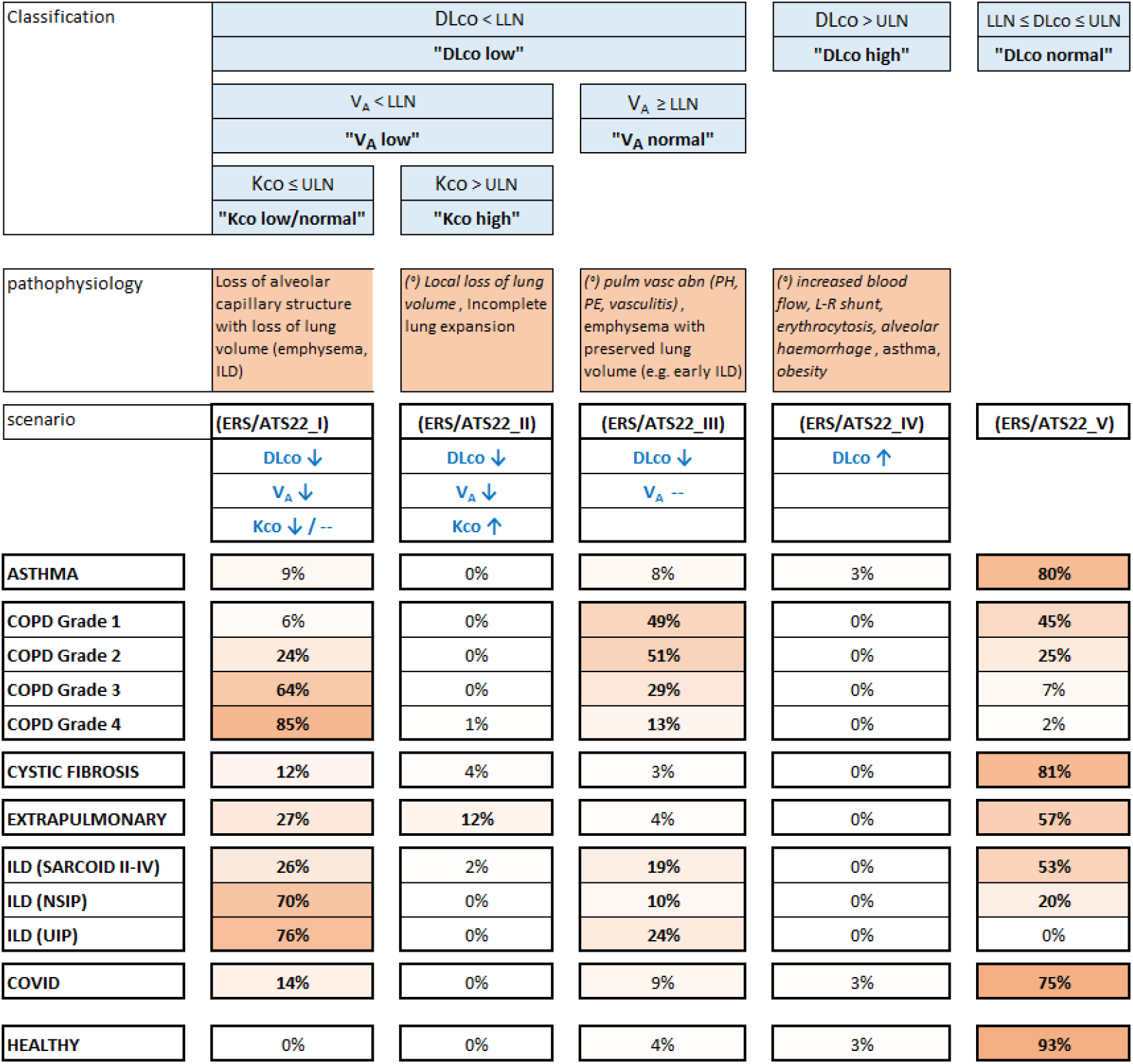
Classification of the six patient groups and the healthy group of Table 1, according to ERS/ATS22 flow chart, interrogating DLco, V_A_ and Kco, and resulting in 5 scenarios, with associated pathophysiological mechanisms. Per subject group (horizontally) all 5 scenarios add up to 100%. Scenarios occurring ≥10% within each group are indicated in bold. Also indicated (blue bold font with arrows) are the variables that were interrogated *(°) italics:* pathologies that could not be explicitly addressed by our patient cohort.

Alternatively, the 9-scenario-flowchart proposed in Figures 2 and 3 (leading to ***1a****-****c, 2a****-****c, 3a****-****c*** with associated pathophysiology) interrogates V_A_ and Kco, where a “low V_A_” is further scrutinized in terms of a patient’s actual lung inflation during the DLco breath-hold phase, TLC_DLco_. TLC_DLco_ is estimated here as plethysmographic residual volume (RV_pl_) plus the inspired volume during the DLco test. V_A_ is considered “low” or “not low” by its V_A_ based z-score threshold at −1.645; TLC_DLco_ is considered “low” or “not low” by its TLC based z-score threshold at −1.645 (using the GLI TLC reference equations for TLC_DLco_). Kco is considered “low” or “normal” or “high” by its Kco based z-score thresholds at −1.645 and +1.645.

**Table 1:** Lung function data for all patients (n= 3474) and healthy subjects (n=303).

|  |  | Asthma<br>n=1615<br>(42% male) |  | COPD<br>n=1338<br>(66% male) |  | Cystic Fibrosis<br>n=108<br>(65% male) |  | Extrapulmonary<br>n=122<br>(46% male) |  | ILD<br>n=98<br>(57% male) |  | COVID<br>n=193<br>(64% male) |  | Healthy<br>n=303<br>(51% male) |  |
| --- | --- | --- | --- | --- | --- | --- | --- | --- | --- | --- | --- | --- | --- | --- | --- |
|  |  | avg | SD | avg | SD | avg | SD | avg | SD | avg | SD | avg | SD | avg | SD |
| age | [years] | 52 | ( 18 ) | 68 | ( 9 ) | 33 | ( 10 ) | 50 | ( 15 ) | 58 | ( 14 ) | 53 | ( 12 ) | 46 | ( 19 ) |
| BMI | [kg/m<sup>2</sup>] | 26 | ( 4 ) | 25 | ( 5 ) | 23 | ( 3 ) | 25 | ( 4 ) | 26 | ( 4 ) | 27 | ( 4 ) | 25 | ( 3 ) |
| packyears |  | 6 | ( 13 ) | 44 | ( 25 ) | 0 | ( 1 ) | 5 | ( 10 ) | 8 | ( 15 ) | 6 | ( 16 ) | 0.4 | ( 1.2 ) |
| FEV<sub>1</sub> | [%pred] | 87 | ( 18 ) | 58 | ( 21 ) | 77 | ( 25 ) | 74 | ( 23 ) | 79 | ( 18 ) | 95 | ( 15 ) | 104 | ( 11 ) |
|  | [z-score] | -0.9 | ( 1.3 ) | -2.5 | ( 1.5 ) | -1.7 | ( 1.8 ) | -1.9 | ( 1.7 ) | -1.4 | ( 1.2 ) | -0.3 | ( 1.0 ) | 0.3 | ( 0.8 ) |
| FVC | [%pred] | 93 | ( 16 ) | 81 | ( 20 ) | 91 | ( 18 ) | 73 | ( 23 ) | 80 | ( 19 ) | 93 | ( 14 ) | 105 | ( 10 ) |
|  | [z-score] | -0.5 | ( 1.1 ) | -1.3 | ( 1.3 ) | -0.7 | ( 1.5 ) | -2.0 | ( 1.8 ) | -1.4 | ( 1.3 ) | -0.5 | ( 1.0 ) | 0.3 | ( 0.7 ) |
| FEV<sub>1</sub>/FVC |  | 0.74 | ( 0.10 ) | 0.55 | ( 0.13 ) | 0.69 | ( 0.15 ) | 0.80 | ( 0.08 ) | 0.78 | ( 0.09 ) | 0.81 | ( 0.08 ) | 0.80 | ( 0.05 ) |
|  | [z-score] | -0.8 | ( 1.2 ) | -2.5 | ( 1.4 ) | -1.8 | ( 1.6 ) | 0.0 | ( 1.1 ) | -0.1 | ( 1.1 ) | 0.3 | ( 1.0 ) | -0.1 | ( 0.7 ) |
| DLco | [%pred] | 93 | ( 18 ) | 58 | ( 21 ) | 91 | ( 18 ) | 78 | ( 18 ) | 68 | ( 22 ) | 89 | ( 19 ) | 102 | ( 13 ) |
|  | [z-score] | -0.5 | ( 1.2 ) | -3.2 | ( 1.8 ) | -0.7 | ( 1.5 ) | -1.7 | ( 1.6 ) | -2.5 | ( 1.9 ) | -0.9 | ( 1.7 ) | 0.1 | ( 0.9 ) |
| Kco | [%pred] | 100 | ( 17 ) | 69 | ( 21 ) | 108 | ( 16 ) | 108 | ( 21 ) | 90 | ( 21 ) | 99 | ( 19 ) | 101 | ( 12 ) |
|  | [z-score] | 0.0 | ( 1.1 ) | -2.2 | ( 1.6 ) | 0.6 | ( 1.2 ) | 0.5 | ( 1.4 ) | -0.7 | ( 1.5 ) | -0.1 | ( 1.4 ) | 0.0 | ( 0.8 ) |
| V<sub>A</sub> | [%pred] | 93 | ( 14 ) | 84 | ( 17 ) | 85 | ( 18 ) | 74 | ( 20 ) | 75 | ( 17 ) | 90 | ( 14 ) | 101 | ( 10 ) |
|  | [z-score] | -0.6 | ( 1.2 ) | -1.4 | ( 1.5 ) | -1.4 | ( 1.8 ) | -2.5 | ( 2.1 ) | -2.3 | ( 1.6 ) | -0.9 | ( 1.2 ) | 0.1 | ( 0.9 ) |
| TLC<sub>pl</sub> | [%pred] | 97 | ( 13 ) | 105 | ( 20 ) | 100 | ( 12 ) | 79 | ( 19 ) | 79 | ( 16 ) | 89 | ( 14 ) | 100 | ( 9 ) |
|  | [z-score] | -0.3 | ( 1.1 ) | 0.4 | ( 1.5 ) | 0.0 | ( 1.0 ) | -1.9 | ( 1.7 ) | -1.8 | ( 1.5 ) | -1.1 | ( 1.5 ) | 0.1 | ( 0.7 ) |
| RV<sub>pl</sub>/TLC<sub>pl</sub> |  | 0.39 | ( 0.11 ) | 0.55 | ( 0.12 ) | 0.36 | ( 0.14 ) | 0.43 | ( 0.11 ) | 0.37 | ( 0.09 ) | 0.35 | ( 0.08 ) | 0.32 | ( 0.08 ) |
|  | [z-score] | 1.3 | ( 1.1 ) | 2.7 | ( 1.7 ) | 2.0 | ( 2.0 ) | 2.1 | ( 1.6 ) | 0.9 | ( 1.0 ) | 0.5 | ( 1.4 ) | 0.6 | ( 0.6 ) |
| V<sub>A</sub>/TLC<sub>pl</sub> |  | 0.89 | ( 0.09 ) | 0.74 | ( 0.15 ) | 0.80 | ( 0.16 ) | 0.86 | ( 0.12 ) | 0.86 | ( 0.07 ) | 0.92 | ( 0.07 ) | 0.92 | ( 0.05 ) |
|  | [z-score] | -0.7 | ( 2.2 ) | -5.1 | ( 5.2 ) | -3.9 | ( 5.8 ) | -1.3 | ( 3.1 ) | -1.1 | ( 1.6 ) | 0.1 | ( 1.6 ) | 0.0 | ( 1.0 ) |

Mean (SD). BMI: body mass index; FEV<sub>1</sub>: forced expiratory volume in 1 s; FVC: forced vital capacity; TLco: transfer factor of the lung for carbon monoxide; Kco: transfer coefficient of the lung for carbon monoxide; V<sub>A</sub>: alveolar volume; RV<sub>pl</sub>, TLC<sub>pl</sub>: residual volume and total lung capacity determined by whole-body plethysmography. GLI was applied to all lung function indices, except for V<sub>A</sub>/TLC, which has no GLI based z-scores. Using V<sub>A</sub>/TLC<sub>pl</sub> data from the healthy group, reference equations were computed by GAMLSS methodology as in our study of reference equations for lung function indices (9,10).

**Figure 2:**
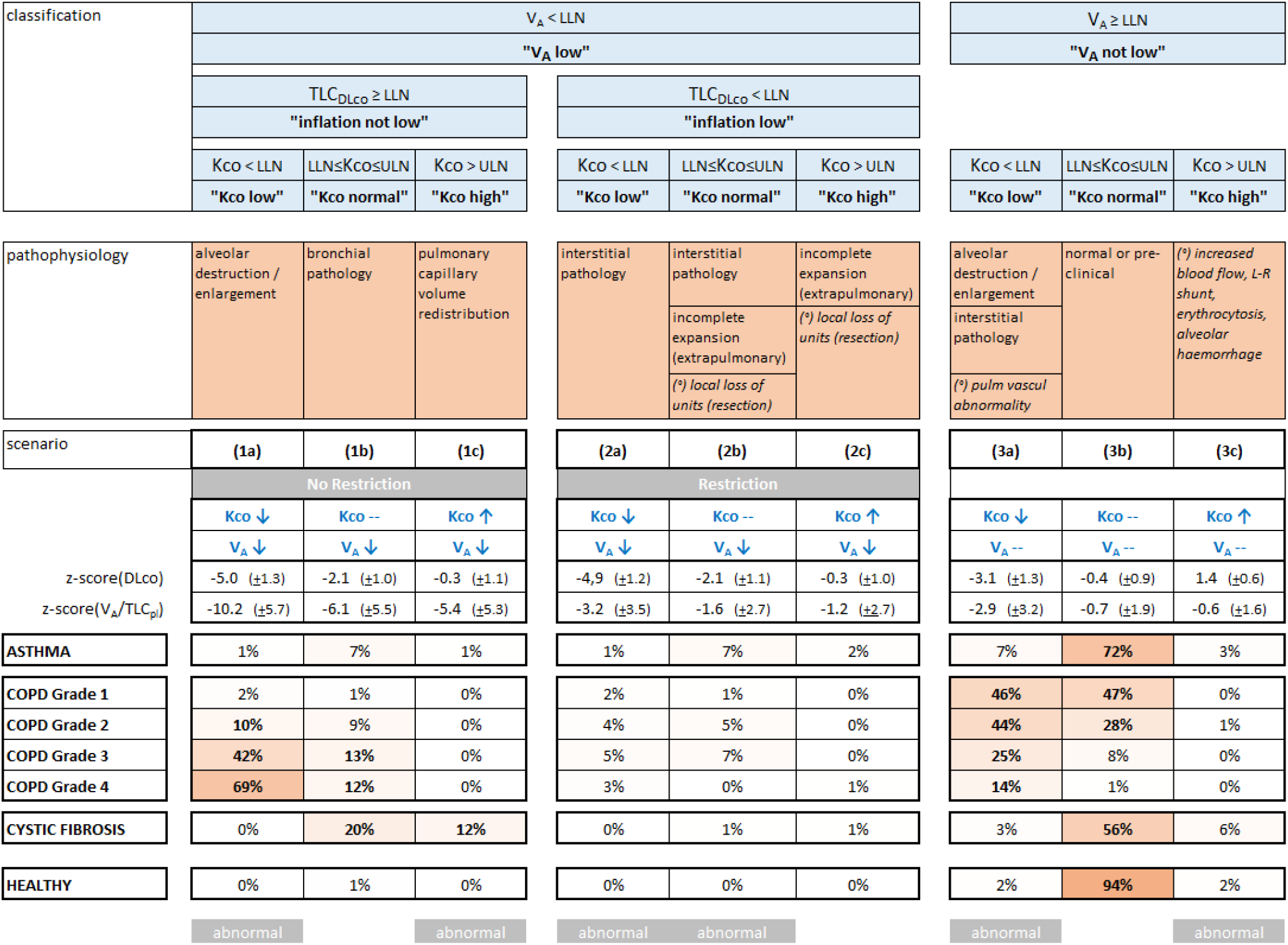
Classification of three patient groups (asthma, COPD, CF) and the healthy group of Table 1, interrogating V_A_ and Kco, and resulting in 9 scenarios, with associated potential pathophysiology. Per subject group (horizontally) all scenarios add up to 100%. Scenarios occurring ≥10% within each group are indicated in bold. Also indicated (blue bold font with arrows) are the variables that were interrogated, and just below are the corresponding mean(+SD) values of DLco_z and V_A_/TLC_pl__z. The scenarios corresponding to an abnormal gas transfer efficiency (abnormal parenchymal and/or pulmonary vasculature) are indicated by grey abnormal boxes. *(°) italics:* pathologies that could not be explicitly addressed by our patient cohort.

**Figure 3:**
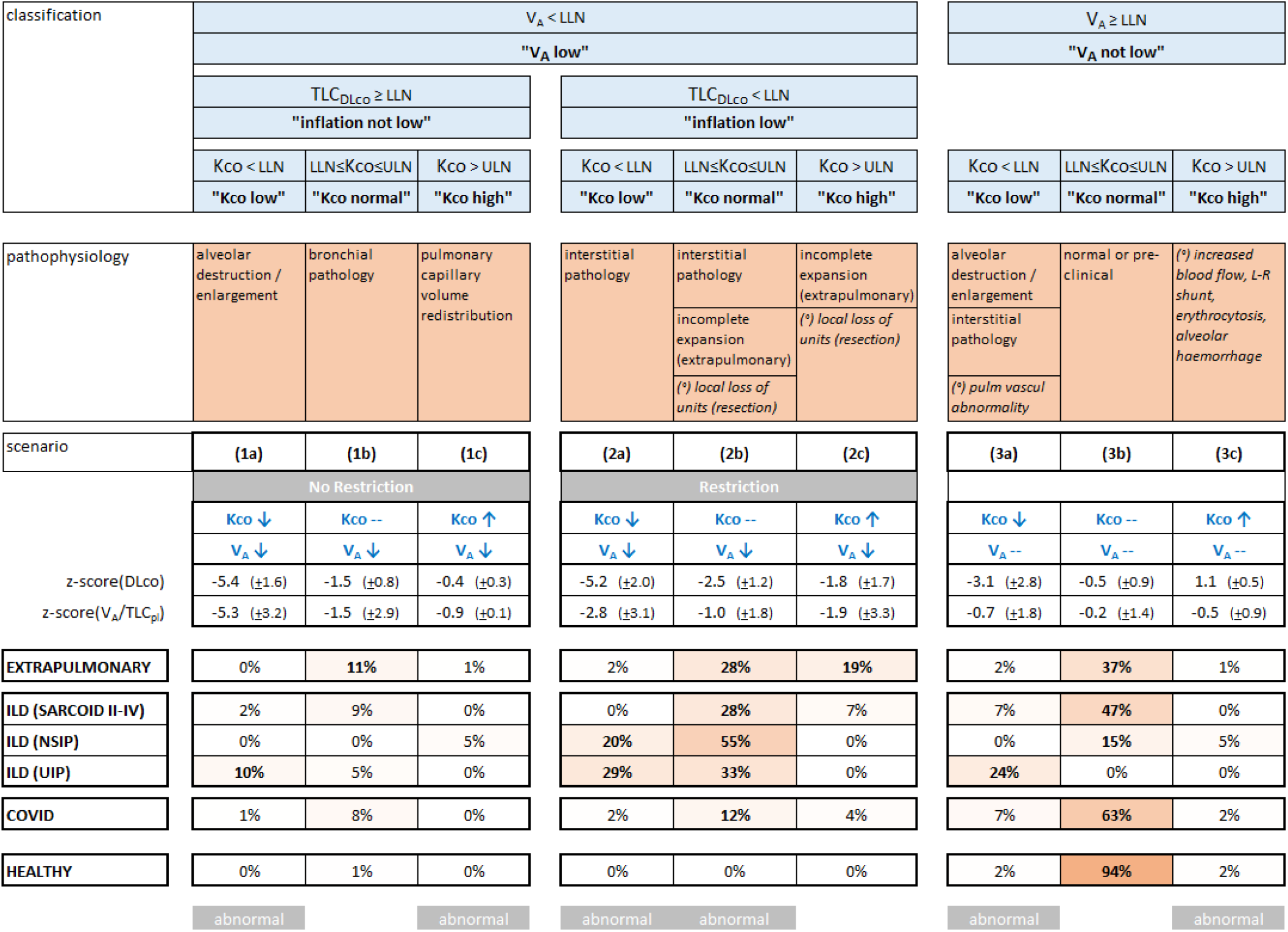
Classification of three patient groups (extrapulmonary restriction, ILD, post-COVID) and the healthy group of Table 1, interrogating V_A_ and Kco, and resulting in 9 scenarios, with associated potential pathophysiology. Same representation as in Figure 2. In the three disease groups combined, z-scores for V_A_/TLC fell below −1.645 in 63% (***2a***), 30% (***2b***) and 46% (***2c***) of patients.

## RESULTS

Three thousand four hundred seventy-four patients could be included for analysis (see flow diagram in OLS1). Table 1 shows pulmonary function for the six clinical diagnosis groups : asthma, COPD (grade 1-4 respectively 16%, 47%, 29%, 9%), cystic fibrosis (CF), disease of extrapulmonary origin, ILD (sarcoidosis II-V, NSIP and UIP respectively 58%, 20%, 21%) and post-COVID.

Figure 1 depicts classification of all patients into 5 scenarios with their corresponding pathophysiology (***I***,***II***,***II***,***IV***,***V***) according to the ERS/ATS22 flowchart; the vast majority of healthy subjects fell into ERS/ATS22_**V**. Figures 2 (asthma, COPD, CF, healthy) and Figure 3 (extrapulmonary, ILD, post-COVID, healthy) classify the same subjects according to the 9-scenario flowchart (**1a-c, 2a-c, 3a-c**) based on the evaluation of V_A_, TLC_DLco_ and K_CO_ ; in this case mean(+SD) z-scores are shown for DLco as the result of this classification. Also shown are V_A_/TLC_pl_ z-scores as an indication of the degree of uneven gas mixing, even though V_A_/TLC_pl_ played no part (was not interrogated) in the 9–scenario classification system itself.

Compared to what is considered a “normal DLco” (ERS/ATS22_***V***; Figure 1), the combination of “normal V_A_” and “normal Kco” in the extended flow chart (***3b***; Figure 2 and 3) obtains similar prevalences in COPD and ILD groups. However, considerable discrepancies arise in asthma, CF, extrapulmonary and post-COVID patient groups, where respectively 8%, 25%, 20% and 11% of patients had an abnormal V_A_ and/or Kco, despite showing a normal DLco.

In Figure 2 and 3, three scenarios (***1b, 2c, 3b***) effectively correspond to a normal gas transfer efficiency : (***1b***) a low V_A_ is entirely due to uneven ventilation and Kco is normal; (***2c***) a low V_A_ is partly due to restriction such that **Kco↑** effectively corresponds to a normal gas transfer (4,6); (***3b***) both V_A_ and Kco are normal. Considering the scenarios corresponding to an abnormal gas transfer efficiency, a patient prevalence of >10% is observed in ***1a*** for COPD (grade 2-4) and UIP, in ***1c*** for adult CF, ***2a*** in NSIP and UIP, ***2b*** in all restricted patient groups of Figure 3, and ***3a*** in UIP.

Figure 4A provides a practical interpretative chart of the likely pathophysiologies associated with each of the 9 V_A_ and Kco scenarios, and Figure 4B illustrates how the 9-scenario flow chart can be readily integrated into routine clinical practice (for a scenario ***1a*** example).

**Figure 4:**
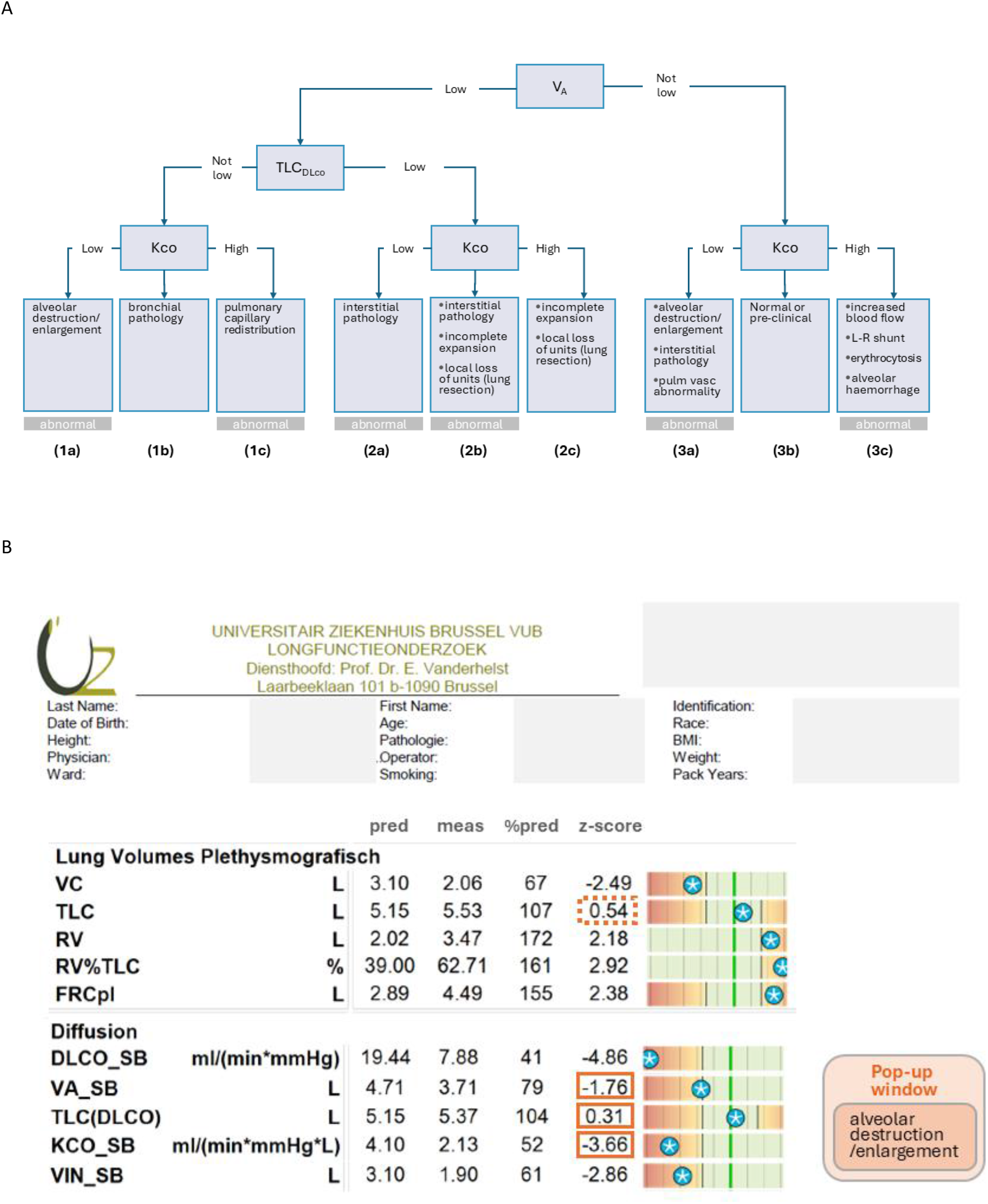
Panel A : Flow chart corresponding to the 9-scenario classification system, adopting the format of Fig.11 in ref 1. The scenarios corresponding to an abnormal gas transfer efficiency (abnormal parenchymal and/or pulmonary vasculature) are indicated by grey abnormal boxes. Panel B : Worked-out example of how the 9-scenario classification can be integrated within routine lung function reporting. Based on 3 z-scores (orange frames) the potential pathophysiology corresponding to scenario ***1a*** in this case, is depicted; the z-score for TLC_DLco_ (=0.31) is obtained by first computing TLC_DLco_ as 5.37 L, 3.47 L (residual volume) plus 1.90 L (inspiratory volume during DLco test) and using the TLC reference values. If expressing TLC_DLco_ in terms of z-score on the lung function report is too difficult, TLC_pl_ z-score could be used instead (orange dotted frame), provided that equipment quality control is sufficient to ensure that inflation reached during the DLco test corresponds to the patient’s TLC obtained during pletysmography.

The Online Supplement shows *(a)* a flow diagram of patient selection; *(b)* raw V_A_/TLC_pl_ values from the 303 healthy subjects and corresponding gamlss based limits of normal for V_A_/TLC_pl_ (Figures OS1,OS2); *(c)* the effect of only considering patients (except COPD) with a smoking history of <5 packyears (Figure OS3).

## DISCUSSION

In this study, we classified 3000+ patients with a variety of lung diseases encountered at a tertiary center into distinct pathophysiological scenarios, depending on the outcomes of the DLco test. Based on the ERS/ATS22 interpretative flowchart, the most prevalent pattern of low DLco was characterized by a low V_A_ and a normal or low Kco (ERS/ATS22_***I***) and contained all lung diseases under study (Figure 1). The predicted occurrence of an abnormally high DLco in asthma (ERS/ATS22_***IV***) could only be observed in 3% of our 1600+ asthma patients.

The first notable difference between the ERS/ATS22 and our novel 9-scenario flow chart was independent of our use of inflation (TLC_DLCO_) to examine a “low V_A_”. Indeed, when just comparing the normal scenarios ERS/ATS22_***V*** (“normal DLco”) and ***3b*** (“normal V_A_” and “normal Kco”), discrepancies up to 25% arise, depending on the lung disease. Of the lung patients under study, CF and extrapulmonary patient groups are seen to benefit most from scrutiny on V_A_ or Kco abnormality beyond a normal DLco.

The full potential of the novel 9-scenario flow chart resulting from assessing a “low V_A_” against lung inflation (TLC_DLCO_) is shown here by the kind of differentiation that can be obtained between patient groups afflicted by obstruction or restriction, or a combination of these (Figure 2,3). These would otherwise find themselves pooled in the same ERS/ATS22 category (Figure 1). The crux of our approach is the identification of a restrictive component, which critically impacts Kco (and resulting DLco) interpretation.

The 9-scenario flow chart highlights two DLco and Kco interpretation paradoxes : (1) patients with abnormal DLco can correspond to normal gas transfer Kco (e.g., 20% of CF patients falling into ***1b***) when low V_A_ is entirely due to uneven ventilation generated by bronchial pathology; (2) patients with normal Kco can in fact correspond to abnormal gas transfer (e.g., 28%-55% of ILD patients falling into ***1b***), because at a low V_A_ due to restriction, Kco should be above normal (assuming reference Kco values are obtained at TLC). The 9-scenario flow chart in Figures 2 and 3, broadly covers effects on DLco from “uneven gas mixing” (scenarios ***1a***-***1c***), “lung restriction” (scenarios ***2a***-***2c***), “membrane or vascular changes” (scenarios ***3a***,***3c***), and “normal or pre-clinical” (scenario ***3b***).

### Uneven gas mixing (scenarios 1a-1c)

A “low V_A_” in the face of a “normal TLC_DLco_” is entirely due to uneven mixing. This was most prominent in COPD and CF patients (Figure 2), but prevalence shifted more towards reduced gas transfer in the more severe COPD patients (**1a**; V_A_↓ Kco↓ thus DLco↓↓). Uneven ventilation as the only contributor to an abnormal DLco test in CF is unsurprising (**1b**; V_A_↓ Kco- - thus DLco↓) given the extended body of evidence on ventilation heterogeneity in CF (13), usually attributed to bronchiectasis. In general, the most likely source of uneven ventilation is bronchial pathology, but emphysema in and of itself (***1a***) can also generate ventilation maldistribution. Uneven ventilation distribution probably also leads to a shift in perfusion distribution towards better ventilated gas exchange units leading to an increase of overall Kco. An interesting scenario in this respect is ***1c*** (V_A_↓ Kco↑ thus DLco--), where the high Kco compensates for the reduced V_A_. The fact that scenario ***1c*** occurs in 12% of CF patients is consistent with enhanced ventilation/perfusion relationship in well-ventilated areas suggested to occur in some CF patients (14).

In cases where low V_A_ is not entirely attributed to uneven mixing (as was the case for ***1a, 1b, 1c***) the V_A_/TLC_pl_ ratio (15) could be used as an indication of ventilation heterogeneity. For instance, in the disease groups with a restrictive component (Figure 3), V_A_/TLC fell below LLN in 63% (***2a***), 30% (***2b***) and 46% (***2c***) of patients. It should be noted that V_A_/TLC_pl_ is a rather crude index of ventilation heterogeneity: age and sex-dependence of V_A_/TLC_pl_ are poor (Figure OLS2) whereas ventilation heterogeneity is known to be age-dependent and increasingly so above ∼50 years (9,16).

### Restricted lung (scenarios 2a-2c)

As expected, low inflation (TLC_DLco_) achievable by patients occurs in ILD, lung disease of extrapulmonary origin or post-COVID (Figure 3). Since Kco is known to be greater at a lung inflation lower than TLC (4,6), expressing Kco in terms of Kco values obtained in normal subjects at normal TLC, implies that in the case of restriction, a Kco between the limits of normal (***2b***) signals an actual *deficit* in gas transfer. This apparent paradox was brought to our attention again during the SARS-CoV-2 crisis where Chapman *et al* (5) laid out the interpretation challenges with the volume dependence of DLco. In our patients studied 10 weeks after a severe SARS-CoV-2 infection, the residual deficit in gas transfer predominantly shows as a Kco between limits of normal (***2b***). This is also the case in 28% of the patients with disease of extrapulmonary origin, which is in line with the observed reduction in dynamic lung compliance in patients with muscle weakness (17), and may reflect subtle abnormalities of the lung parenchyma or pulmonary vasculature in these patients. When focusing on the ILD patients, the UIP and NSIP subgroups show a shift from a moderate gas transfer deficit (***2b***) towards a more marked one (***2a***), which could be expected based on their respective disease profile (18).

### Membrane or capillary vascular changes (scenarios 3a,3c)

Where V_A_ was normal, an isolated low Kco (***3a***; V_A_--Kco↓ thus DLco↓) was primarily observed in the low COPD grades and in UIP, probably reflecting early disease stages where V_A_ is not yet affected (by uneven gas mixing and/or restriction). While the ***3a*** scenario is also a marker for pulmonary vascular disease, such patients were not seen amongst our pulmonary function test referrals (see ***Limitations***). An isolated high Kco (***3c***; V_A_--Kco↑ thus DLco↑) was unusual in the patients under study, unless in 6% of CF patients likely displaying areas of high perfusion, similar to the above-discussed case of (***1c***) but without the impact of uneven mixing on V_A_.

### Normal or pre-clinical (scenario 3b)

The pattern where all constituent parts of DLco are normal (V_A_--Kco--thus DLco--) was observed in more than half of the patients in asthma (72%), post-COVID (63%), and CF (56%), followed by sizeable portions of patients with grade 1 COPD (47%), sarcoidosis II-IV (47%) and lung disease of extrapulmonary origin (37%) (Figure 2, 3). While it is easy to accept that V_A_ and Kco can be normal in asthma, it may be more surprising in other disease groups. This could simply reflect a lack of sensitivity of standard lung function tests.

### Scrutinizing V_A_ against actual lung inflation

Lung inflation TLC_DLCO_ is computed here as pletysmographic RV plus inspired volume during the DLco test, whereas RV can also be determined from rebreathing or multiple breath washout techniques. If TLC_DLCO_ is too difficult to integrate with the lung function report (Figure 4B), one could even consider using TLC_pl_ as a surrogate. However, if no reliable TLC estimate is available at all, this would imply a pairwise collapse of (***1a***,***2a***), (***1b***,***2b***) and (***1c***,***2c***). As a result, patients with normal gas transfer efficiency (***1b***), for instance 20% of the CF patients, would get pooled with patients with a restrictive component who do have parenchymal or vascular abnormalities (***2b***).

### Clinical application

Now that all age GLI reference values exist for DLco components Kco and V_A_ (11), and lung volumes RV and TLC (12), the flowchart of Figure 3A can be easily integrated within current lung function reporting as shown in Figure 3B. It can potentially also be applied in the pediatric range, for instance in the lung function workup of ex-preterm children, where uneven ventilation, restriction and impaired alveolar-capillary membrane abnormalities may contribute to DLco (19). The 9-scenario flowchart could also inform AI based classification systems such as the one described by Desbordes *et al* (20) where the authors acknowledged as a study limitation that alveolar volume was not used.

### Limitations

Some conditions mentioned in the ERS/ATS2022 guideline document could not be investigated, such as pulmonary hypertension or embolism (***3a***), left-right shunt, erythrocytosis or alveolar hemorrhage (all ***3c***) because of a lack of referrals of such patients to the pulmonary function laboratory.

***In conclusion***, this is the first study to classify a large and varied patient cohort in terms of their DLco characteristics based on the ERS/ATS 2022 chart, as well as an extended 9-scenario classification system where V_A_ is assessed against actual lung inflation during the DLco test. This novel classification expands the interpretative capabilities and potentially brings more nuance to the various etiologies of an abnormal pulmonary diffusion test in the clinic.

## Supporting information

Supplement to 20260130_Verbanck_MainText.pdf

## Data Availability

All data produced in the present study are available upon reasonable request to the authors.

## Author Contributions

S.V(e)., M.H. conceived of the study and drafted the manuscript; S.V(e)., F.D., S.W., S.V(i)., E.V., S.H. analyzed the data; all authors revised the manuscript.

