## Supplement to 20260130_Verbanck_MainText.pdf for "Appraisal and extension of the ERS/ATS Interpretative Strategy for Pulmonary Diffusing Capacity"

1. <u>Flow chart of subject selection in this study</u>

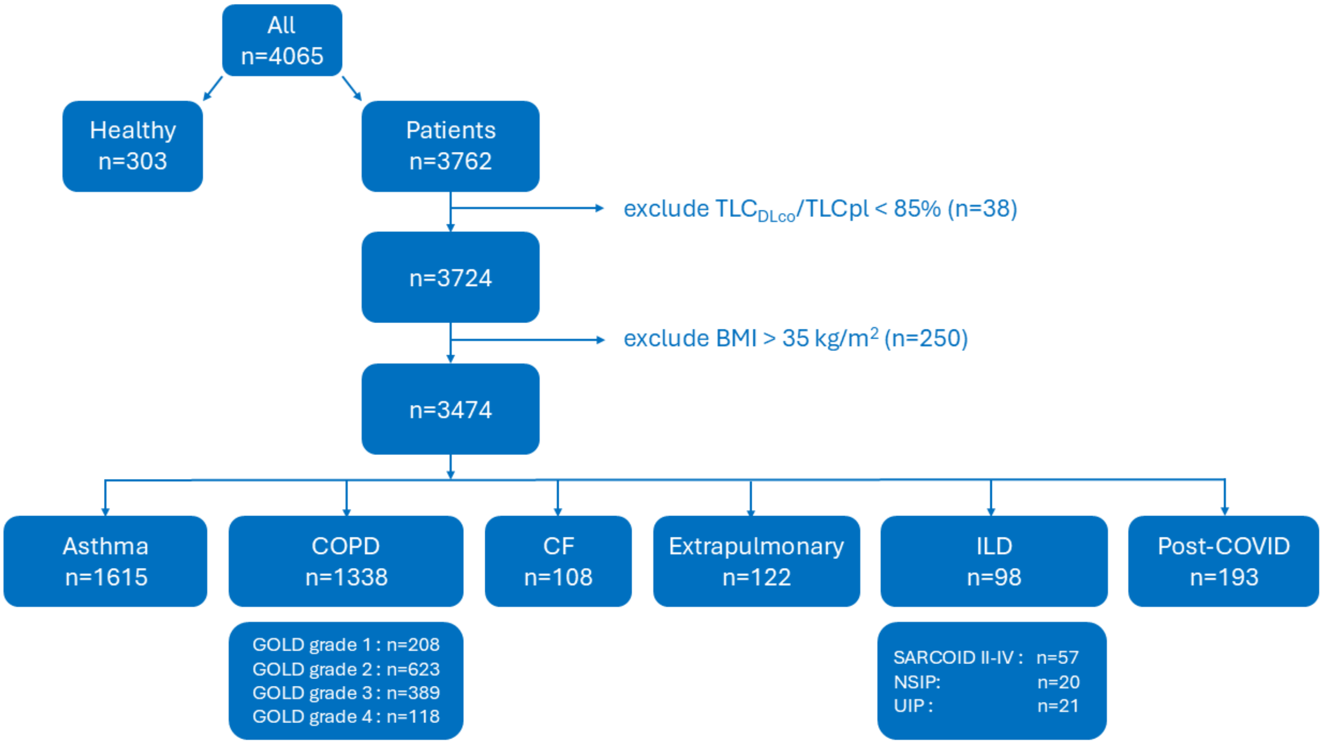


Abbreviations : CF : cystic fibrosis; ILD : interstitial Lung disease; Post-COVID : patients at 10 weeks after having suffered a severe SARS-CoV-2 infection; TLCpl, $TLC_{DLCO}$ : total lung capacity from pletysmography, TLC during the DLco test, i.e., the sum of $RV_{pl}$ and inspired capacity.

2. Quality control for included DLco tests

All measurements were done on lung function equipment (MasterScreenPFT, SentrySuite, Mettawa, IL,USA) in accordance with current quality standards (refOLS1, refOLS2). Only “good quality” tests (as per equipment specifications) were retained, and complemented with anthropometric data and pneumologist diagnosis, before sending these to the local database (SentrySuite CIS Datacube 3.20). In addition to the equipment’s standard quality control measures regarding inspired capacity (above 85-90% of a known vital capacity), we further verified that only DLco data from those maneuvers where lung inflation exceeded 85% of the patient’s total lung capacity ($TLC_{pl}$) were accepted as valid for further analysis. This was done by establishing the patient’s actual lung inflation during the DLco breath-hold phase ($TLC_{DLco}$) as plethysmographic residual volume ($RV_{pl}$) plus the inspired volume during the DLco test. When $TLC_{DLco}$ was greater than 85% of the patient’s plethysmographic TLC ($TLC_{pl}$) DLco tests were considered valid.

3. Normal values of $V_A$, TLC and $V_A$/TLC

Using $V_A/TLC_{pl}$ data from the healthy subjects, reference equations were computed using the same methodology as in our study of reference equations for lung function indices (9,10) (GAMLSS package; version2.15.2; R-Foundation, Vienna, Austria).

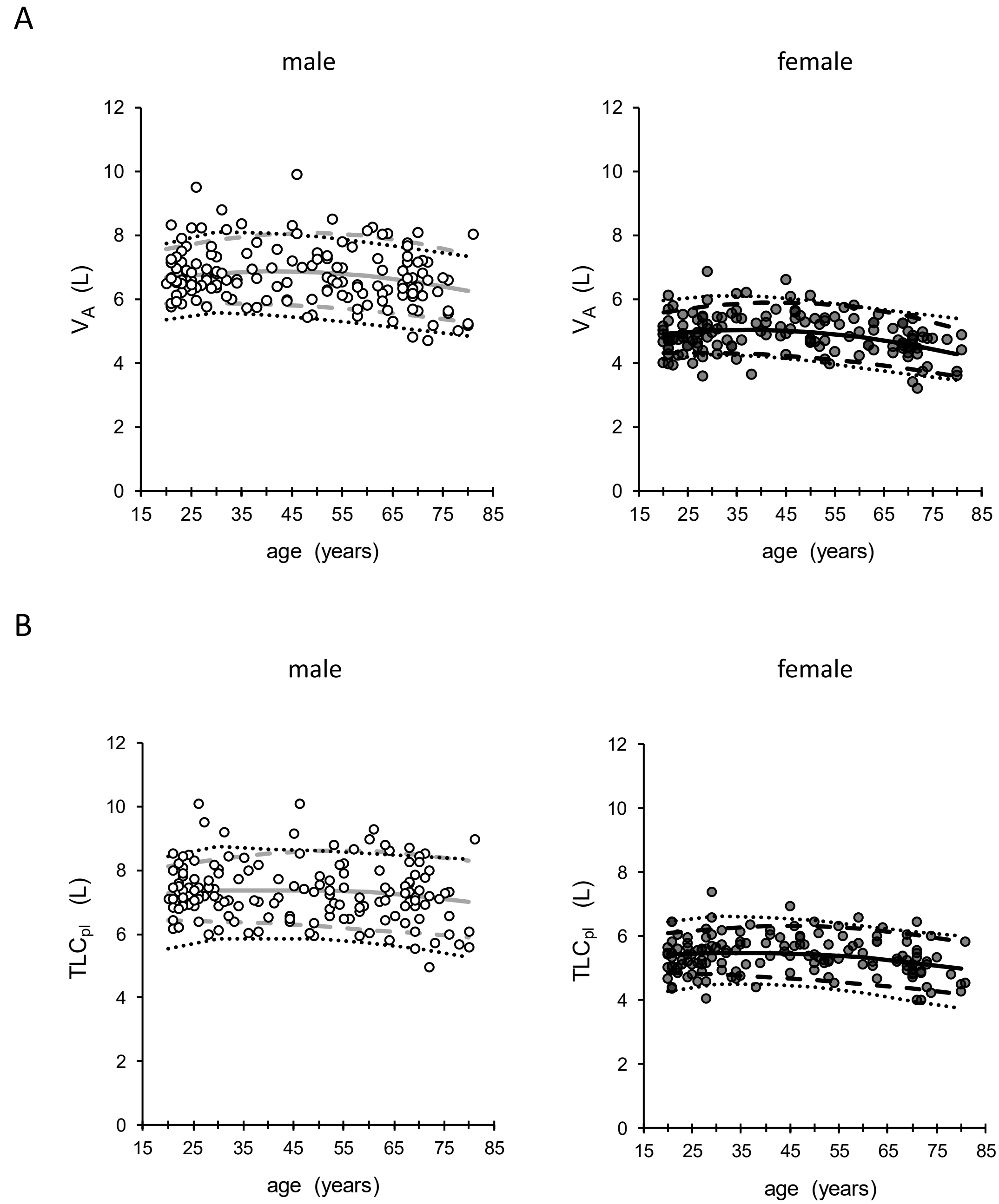


Figure OS1 : Scatterplots of $V_A$ (panel A) and $TLC_{pl}$ (panel B) obtained on all healthy subjects in the 20-80 age range (n=303); male : open circles and grey lines for median, lower and upper limits of normal (LLN, ULN); female: solid circles and black lines for median, LLN and ULN; dotted lines are the corresponding ULN and LLN obtained from GLI (https://gli-calculator.ersnet.org/ last accessed 25 August 2025). The age dependence of median, LLN and ULN curves also takes into account the height dependence with age in this group.

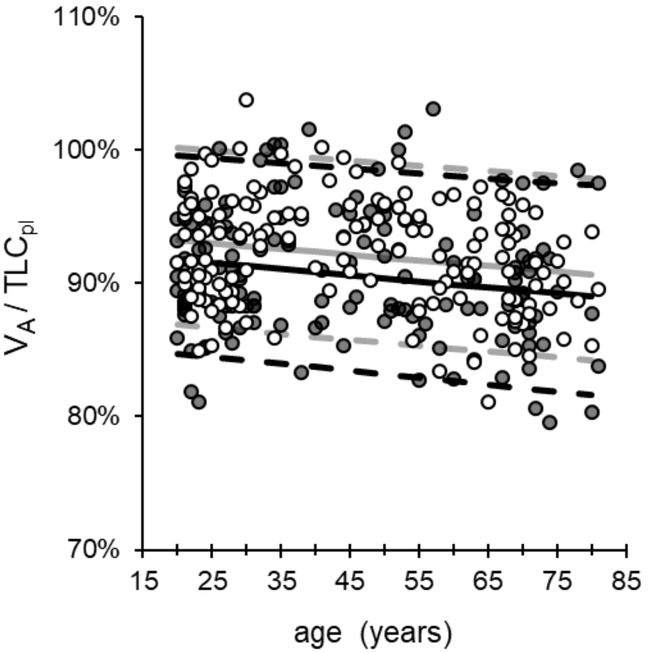


Figure OS2 : Scatterplots of $V_A/TLC_{pl}$ obtained on all healthy subjects in the 20-80 age range (n=303); male : open circles and grey lines for median, lower and upper limits of normal (LLN, ULN); female: solid circles and black lines for median, LLN and ULN. The age dependence of median, LLN and ULN curves also takes into account the height dependence with age in this group. Overall, these LLN compare well with the constant LLN of 83% obtained by Roberts *et al* (15) for normal subjects (M/F) aged 18-86.

TABLE OS1 : The M, L and S coefficients to obtain predicted values and lower and upper limits of normal for alveolar volume ($V_A$), total lung capacity ($TLC_{pl}$) and $V_A/TLC_{pl}$.

| | | M coefficients | | | | | S coefficients | | | | L coefficient | |
|---|---|---|---|---|---|---|---|---|---|---|---|---|
| | | sex | height | age | age$^2$ | intercept | height | age | age$^2$ | intercept | intercept | $r^2$ |
| $V_A$ | [ L ] | -1.021 | 0.0659 | 0.0392 | -0.000412 | -5.81 | -0.00184 | 0.01934 | -0.00015 | -2.55 | 0.233 | 0.78 |
| $TLC_{pl}$ | [ L ] | -1.029 | 0.0704 | 0.0263 | -0.000251 | -5.86 | 0.00291 | 0.02031 | -0.00014 | -3.52 | 0.427 | 0.79 |
| $V_A$ / $TLC_{pl}$ | | -0.010 | 0.0005 | -0.0004 | | 0.86 | -0.01098 | 0.00000 | | -1.16 | -0.358 | 0.08 |

### 4. Effect of smoking history.

| classification | $V_A$ < LLN "$V_A$ low" | | | | | | $V_A$ ≥ LLN "$V_A$ not low" | | | |
|---|---|---|---|---|---|---|---|---|---|---|
| | $TLC_{DLco}$ ≥ LLN "inflation not low" | | | $TLC_{DLco}$ < LLN "inflation low" | | | | | | |
| | Kco < LLN **"Kco low"** | LLN≤Kco≤ULN **"Kco normal"** | Kco > ULN **"Kco high"** | Kco < LLN **"Kco low"** | LLN≤Kco≤ULN **"Kco normal"** | Kco > ULN **"Kco high"** | Kco < LLN **"Kco low"** | LLN≤Kco≤ULN **"Kco normal"** | Kco > ULN **"Kco high"** | |
| pathophysiology | alveolar destruction / enlargement | bronchial pathology | pulmonary capillary volume redistribution | interstitial pathology | interstitial pathology<br>incomplete expansion (extrapulmonary)<br>*(°) local loss of units (resection)* | incomplete expansion (extrapulmonary)<br>*(°) local loss of units (resection)* | alveolar destruction / enlargement<br>interstitial pathology<br>*(°) pulm vascul abnormality* | normal or pre-clinical | *(°) increased blood flow, L-R shunt, erythrocytosis, alveolar haemorrhage* | |
| scenario | **(1a)** | **(1b)** | **(1c)** | **(2a)** | **(2b)** | **(2c)** | **(3a)** | **(3b)** | **(3c)** | % patients with <5py |
| **ASTHMA** | 1% | 7% | 1% | 0% | 8% | 2% | 5% | **72%** | 3% | 75% |
| **COPD Grade 1** | | | | | | | | | | |
| **COPD Grade 2** | | | | | | | | | | |
| **COPD Grade 3** | | | | | | | | | | |
| **COPD Grade 4** | | | | | | | | | | |
| **CYSTIC FIBROSIS** | 0% | **21%** | **13%** | 0% | 1% | 1% | 3% | **55%** | 7% | 96% |
| **EXTRAPULMONARY** | 0% | **12%** | 0% | 1% | **27%** | **18%** | 1% | **39%** | 1% | 75% |
| **ILD (SARCOID II-IV)** | 0% | 8% | 0% | 0% | **30%** | 4% | 6% | **52%** | 0% | 88% |
| **ILD (NSIP)** | 0% | 0% | **13%** | **13%** | **63%** | 0% | 0% | **13%** | 0% | 40% |
| **ILD (UIP)** | **13%** | **13%** | 0% | **13%** | **50%** | 0% | **13%** | 0% | 0% | 38% |
| **COVID** | 0% | 9% | 0% | 2% | **13%** | 5% | 5% | **64%** | 3% | 77% |

Figure OS3 : Equivalent of Figure 2 and 3, but by only including patients with a less than 5 packyears smoking history, considering all patient groups except for COPD.
